# An Occupational Health Survey for Port Employees in Shenzhen and A Dataset Management System based on R

**DOI:** 10.1101/2022.06.27.22276896

**Authors:** Jinlin Wang, Chunbao Mo, Yuehong Huang, Dafeng Lin, Jie Situ, Ming Zhang, Naixing Zhang

## Abstract

**Background:** Port employees is a huge occupational group in industrial economy but the occupational health problem does not receive enough attention and the relative data is still deficient. Hence, the aim of the study was to survey the occupational health condition for port employees and to construct a relative dataset.

**Method:** A cross-sectional study was implemented among the population in a Shenzhen port, they were required to undergo occupational physical examination and questionnaires to learn about physical condition and other information. Description analysis were used to describe the data, and missing value analysis and Cronbach’s alpha coefficient were utilized to evaluated the data quality. And management system based on Shiny was constructed to manage and analyze the dataset.

**Result:** A total of 5245 participants involved in this study, 3211 of them received occupational physical examinations, 3946 participants received the questionnaire, and 1912 received the both. Quality analysis suggested that the total missing rate of these three datasets were 10.76%, 0% and 4.78%, respectively. And the total Cronbach’s alpha confidence of Effort-Reward Imbalance Questionnaire and National Health Literacy Monitoring Questionnaire was 0.808. Furthermore, a dataset management system with preview overview, selection, output and summary functions was constructed.

**Conclusion:** Occupational Health Survey for Port Employees is a reliable survey and it’s system can be used to manage and analyze the dataset, however, further optimization and improvement are still required.

## Introduction

The seaport plays an increasingly critical role as the center of ocean transportation, with the development of international trade^1, 2^. According to statistics, there were 1.2 million people currently working at sea in the world in 2009^3^. Among the European Union’s 22 maritime member states, over 110,000 port workers in the loading and unloading of ships^4^. In China, a big maritime trading nation, there are over 808,000 maritime employees^5^. Because of the complex process, port employees usually exposure to various occupational hazards which may lead to accidental injury or occupational diseases, for instance, ergonomic hazards (e.g., repetition of movements, awkward positions), physical hazards (e.g., noise, vibration, heat, radiation), biological agents (e.g., virus, bacteria, insect), chemical substances and psychosocial hazards (e.g., overload, stress, shift work)^6^. Even with technological advances and numerous mechanical equipment are utilized, abundantly manual labor is still required. Therefore, it is necessary to pay attention to the occupational health for the port employees who exposure to occupational hazards.

Several observational epidemiology research were implemented to develop new knowledge in the field of occupational safety and health and to transfer that knowledge into practice. The National Institute for Occupational Safety and Health (NIOSH), a part of the U.S. Centers for Disease Control and Prevention, established a huge and extensive occupational health database via epidemiology investigation for occupational populations, which was useful for exploring the factors affecting occupational health and proposing protection strategies, such as Occupational Hearing Loss Surveillance Project (OHLSP)^7^ and The National Study of Coal Workers’ Pneumoconiosis (NSCWP)^8^. And the National Institute for Occupational Health (NIOH) in South Africa has an established research record of more than 60 years involving in bio-aerosol exposure in the workplace, dusts and dust related diseases, ergonomics, reproductive health and other^9, 10^. They were long term research projects and mostly focused on coal mining, commodity production, or building industry. For the port employees, although a few occupational health studies had been reported, the research attention and relative data is still deficient. Similarly, in China, absence still exist in this field, so it is necessary to strengthen the research on occupational health of port employees and establish a corresponding research database.

How to manage and utilize data efficiently is a basis question needing researchers to answer. On the one hand, database management needs the support of embedded software provided by a third-party, which needs to pay purchase or lease fee. On the other hand, extra professionals are needed to maintenance and upgrade this software. Moreover, it will be a risk for data loss or leakage relying on the data storage and management services provide by a third-party. Hence, developing a private, highly customized database management system by a simple, reliable, low-cost approach may be a reliable option. With free and open-source programming techniques becoming popular, programming is no longer the specialty of professional programmers, researchers have mastered at least one programming language, such as R or Python in increasing numbers^11, 12^. R is a free software environment for statistical computing and graphics, and the Shiny is an R package that makes it easy to build interactive web applications directly^11, 12^, so in this study, we tend to build a database management system based on Shiny package to achieve data management, analysis and visualization.

Shenzhen, one of the most economically developed cities in China, located in the south of Guangdong Province with two advanced ports and tens of thousands of employees. It is a good experimental place for researching occupational exposure and health. In 2020, we conducted an epidemiological survey for port employees in Shenzhen and constructed a dataset management system based on Shiny, aiming at understanding population health condition, identifying occupational harmful factors and providing basic data for protection policy formulation.

## 2. Method

### 2.1 Study design and participants

Occupational Health Survey for Port Employees (OHSPE) was an occupational hygiene research conducted by Shenzhen Prevention and Treatment Center for Occupational Diseases (SPTCOD) for port employees in Shenzhen, aiming to know about the health condition, evaluate occupational hazards exposure and explore the influencing factors for port employees. From June to October 2020, an occupational health examination among port employees in Shenzhen was implemented with a cross-sectional design, followed by a questionnaire several months later. The inclusion criteria were 1) the participant was employed in a port of Shenzhen, 2) who worked more than one year and 3) understood the purpose and significance of the study and signed the informed consent voluntarily. The exclusion criteria were 1) non-port workers, 2) not cooperating with investigators. Participants were re-assigned unique identifier number which associated with one’s case report forms (CRFs), study reports and biospecimen. This study complied with the Helsinki Declaration and had been approved by Medical Ethics Committee of SPTCOD (NO. LL2020-34).

### 2.2 Occupational physical examinations

Participants were required to undergo a routine physical examination, chest CT, abdominal ultrasound, ophthalmology, ear and motor nerve function tests by professional physicians to collect anthropometric (such as height, weight, blood pressure, etc.), lung, abdominal organs, vision, hearing and other indicators related to health. Urine and elbow venous blood samples were collected and sent to the laboratory for testing biochemical indicators including determination of fasting blood glucose, total cholesterol (TC), high-density lipoprotein cholesterol (HDL-C), low-density lipoprotein cholesterol (LDL-C), triiodothyronine (T3), thyroxine (T4), etc., by using automatic biochemical analyzer. The above examination and testing were implemented in SPTCOD.

### 2.3 Questionnaires

Structured electronic questionnaires were distributed to the participants by the uniformly trained investigators and filled in by themselves. The questionnaire consisted of the following four parts: 1) baseline information: name, gender, ID number, nation, education level, job, length of service, working hours per week, working environment, etc.; 2) Lifestyle and health condition: smoking, alcohol consumption, medical history according to physician diagnosis and physical activity; 3) Occupational stress assessment: The Effort-Reward Imbalance Questionnaire (ERIQ) based on the Effort-Reward Imbalance theoretical model was utilized to investigates job stress through the imbalance between the efforts the worker makes (costs) and the rewards he or she receives (gains), this questionnaire contained three dimensions of effort, return and overload, with 6, 11 and 6 items respectively, and it had been applied widely and proved to have good reliability and validity for its Chinese version^14, 15^; 4) Health literacy assessment: According to the expert advice and the traits of jobs of participants, the National Health Literacy Monitoring Questionnaire (2014 edition, NHLMQ) ^16, 17^was amended to adapted the investigation for evaluating the health literacy of participants in this study. This specialized questionnaire consisted of four dimensions of healthy knowledge, attitude, behavior and skills, with 64 items.

### 2.4. Construction of information management system

Two R scripts were needed to construct a dataset management system, the first one is ui.R, which was utilized to build user-facing graphical interfaces and the second one is server.R, it was a sever script to execute command. This study would adopt the Dashboard design to build a dataset management system with functions of query, summary, screening, description, analysis and visualization.

### 2.5 Statistical method

Descriptive analysis was used to report and describe the main data of this survey, continuous variables were expressed using the mean ± standard deviation (*x*□±s), whereas categorical variables were described using the number of cases and composition. Cronbach’s alpha coefficients could be utilized to evaluate the reliable for questionnaire, the good Cronbach’s alpha coefficients were set to be 0.8 or greater, which indicates the internal consistency of this survey is well^18^. The dataset management system would be developed by using R software and its packages, such as “Shiny”, “shinyWidgets”, “Tidyverse”, “DT”, “HighCharter” and “ShinyDashboard”.

## 3. Result

### 3.1 Characteristics of the study population

There were 5245 participants involved in this study, 3211 of them received occupational physical examinations (examinations set), and 3946 participants received the questionnaire (questionnaire set), however only 1912 received the both (both set). In these subjects, most of them were male, accounting for more than 95%, and the average age was about 40 years old, and the majority nation was Han. The education level was dominated by those who had received junior or senior high school, and followed by college degree or above. In the questionnaire set and both set, the mean length of service was 15.2 and15.9 years, respectively. For descripting the result conveniently, these jobs were divided into the following categories, because of the numerous stall positions in the port. 1) Driver, refers to a person who drives or operates a vehicle, forklift, loader, crane, etc.; 2) Longshoremen; 3) Skilled worker, including electrician, welder, maintenance, etc.; 4) Management including manager, executives, etc.; 5) Security, including guard and patrol 6) Inspector, including tallyman and receiving clerk; 7) Clerk including accountant and telephone operator; and 8) other jobs, for example handyman, pilot, etc. The percentages of jobs varied in different sets, in examinations set and both set the jobs distribution was the same, the top three were driver, longshoremen and skilled worker. However, in questionnaire set the top three were other jobs, longshoremen and skilled worker, table 1.

**Table 1.** The demographic characteristic for port employees in this study

|  | Items | Examinations set<br>N=3211 | Questionnaire set<br>N=3946 | Both set<br>N=1912 |
| --- | --- | --- | --- | --- |
| Gender | Male | 3075 (95.8%) | 3756 (95.2%) | 1840 (96.2%) |
|  | Female | 136 (4.2%) | 190 (4.8%) | 72 (3.8%) |
| Age | Mean (SD) | 39.6 (11.1) | 40.0 (10.2) | 40.3 (10.9) |
|  | Median (min, max) | 40.0 (18.0, 64.0) | 40.0 (18.0, 60.0) | 41.0 (18.0, 59.0) |
| Nation | Han | - | 3750 (95.0%) | 1809 (94.6%) |
|  | Other | - | 196 (5.0%) | 103 (5.4%) |
| Marriage | Yes | 2234 (69.6%) | 2903 (73.6%) | 1415 (74.0%) |
|  | No | 634 (19.7%) | 905 (22.9%) | 433 (22.6%) |
|  | Other | 343(10.7%) | 138 (3.5%) | 64 (3.3%) |
| Education level | Primary school or below | - | 195 (4.9%) | 107 (5.6%) |
|  | Middle or high schools | - | 2533 (64.2%) | 1504 (78.7%) |
|  | College degree or above | - | 1218 (30.9%) | 301 (15.7%) |
| Length of service | Mean ( SD ) | - | 15.2 (9.75) | 15.9 (10.3) |
|  | Median (min, max) | - | 15.0 (1.00, 49.0) | 15.0 (1.00, 44.0) |
| Jobs | Driver | 1752 (54.6%) | 495 (12.5%) | 918 (48.0%) |
|  | Longshoremen | 381 (11.9%) | 1041 (26.4%) | 274 (14.3%) |
|  | Skilled worker | 283 (8.8%) | 529 (13.4%) | 254 (13.3%) |
|  | Management | 248 (7.7%) | - | 185 (9.7%) |
|  | Security | 220 (6.9%) | 119 (3.0%) | 112 (5.9%) |
|  | Inspector | 173 (5.4%) | 258 (6.5%) | 95 (5.0%) |
|  | Clerk | 38 (1.2%) | - | 14 (0.7%) |
|  | Other jobs | 116 (3.6%) | 1504 (38.1%) | 60 (3.1%) |

### 3.2 Data quality

The total Cronbach’s alpha confidence of ERIQ and NHLMQ was 0.808, 95% confidence interval: 0.788-0.827, which indicated that the questionnaire was reliable. Table 2 exhibited the result of missing value analysis. No missing value were existed in questionnaire set. But missing value were observed in examination set and both set, the overall missing rate were 10.76% and 4.78%, respectively. Furthermore, in the examination set the mean missing rate of column (i.e., physical examination items) was 10.76%, with the range of 100%∼0%. And the mean missing rate of row (i.e., sample items) was 10.76%, with the range of 8.89% ∼12.58%. In summary, the quality of examination dataset was acceptable. There were a few missing values in the both set, the missing rate of both column and row was 4.78%, which suggested the quality was good. On the contrary,

**Table 2.** result of missing value analysis

|  | Items | Examinations set | Questionnaire set | Both set |
| --- | --- | --- | --- | --- |
| Dimension | Column | 167 | 205 | 371 |
|  | Row | 3211 | 3946 | 1912 |
| Overall missing | Number | 57682 | 0 | 33876 |
|  | Rate (%) | 10.76 | 0 | 4.78 |
| Column missing rate | Mean (SD, %) | 10.76 (30.29) | 0 | 4.78 (20.75) |
|  | Median (min, max, %) | 0 (0,100) | 0 | 0 (0,100) |
| Row missing rate | Mean (SD, %) | 10.76 (0.90) | 0 | 4.78 (0.46) |
|  | Median (min, max, %) | 11.38 (8.89, 12.58) | 0 | 4.58 (4.04, 5.66) |

### 3.3 Dataset management system

Basing on Shiny package, a dataset management system was be constructed: the Occupational Health Survey for Port Employees Information Management System (OHSPEIMS), which home page and structure were exhibited in Figure 1. On the sidebar of OHSPEIMS contained four menus of Home, Dataset, Analysis and About. The main interface and basic information of hosting institution, researchers and acknowledgement about OHSEP project were provide on the Home and About pages, respectively. In addition, Dataset option provides three submenus, showing Baseline Data, Physical and Examination Data and Questionnaire Data of the samples in different interfaces respectively, and providing data preview overview, selection, output and summary functions (Fig2 A-B). In Analysis option, Exploratory Analysis module provided chi-square test, T test, line regression and logistic regression models to perform exploratory or correlation analysis (Fig2 C). And lotting Scatter diagram and fitting curve or other graphs could be achieved in the Plotting module (Fig2 D).

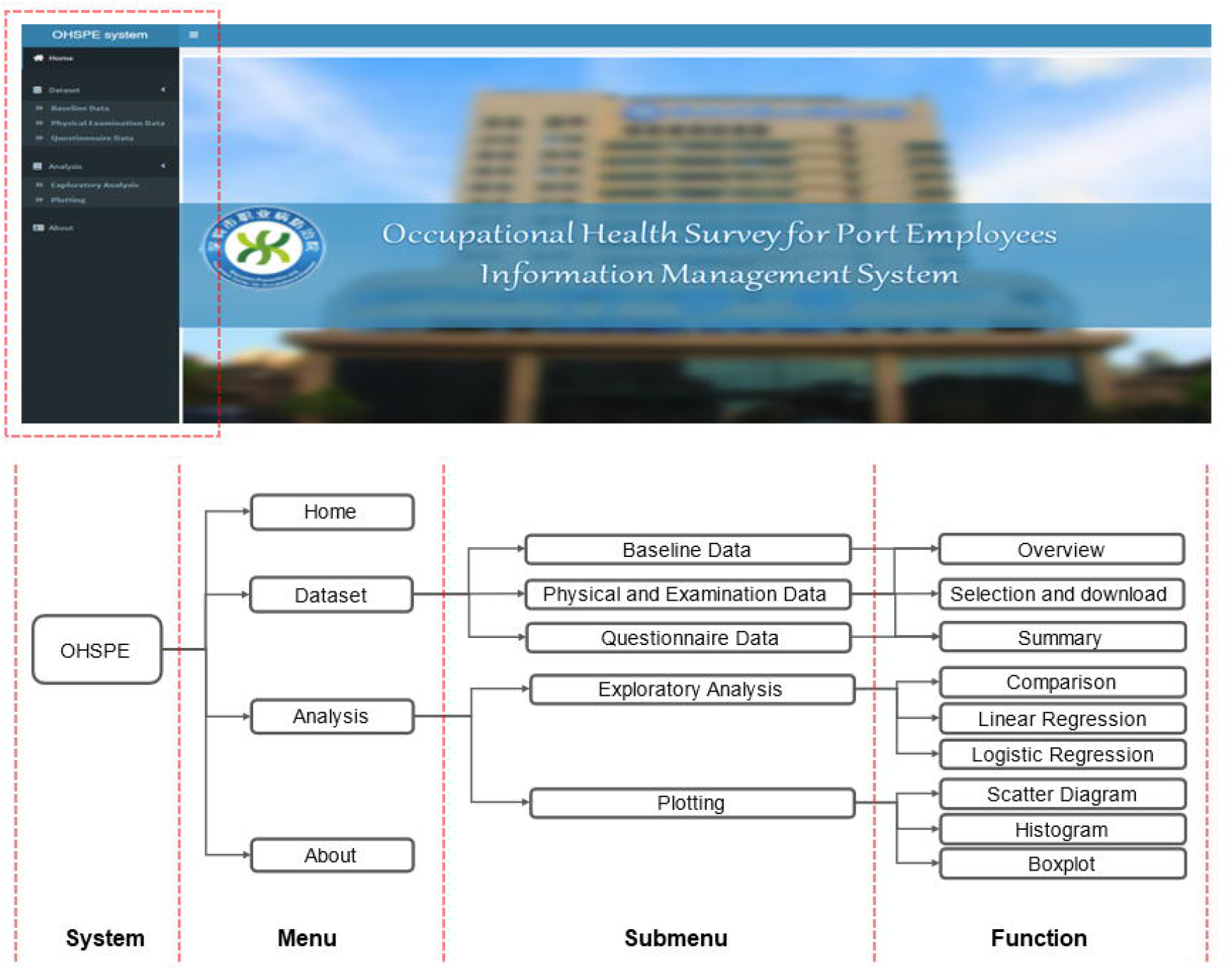

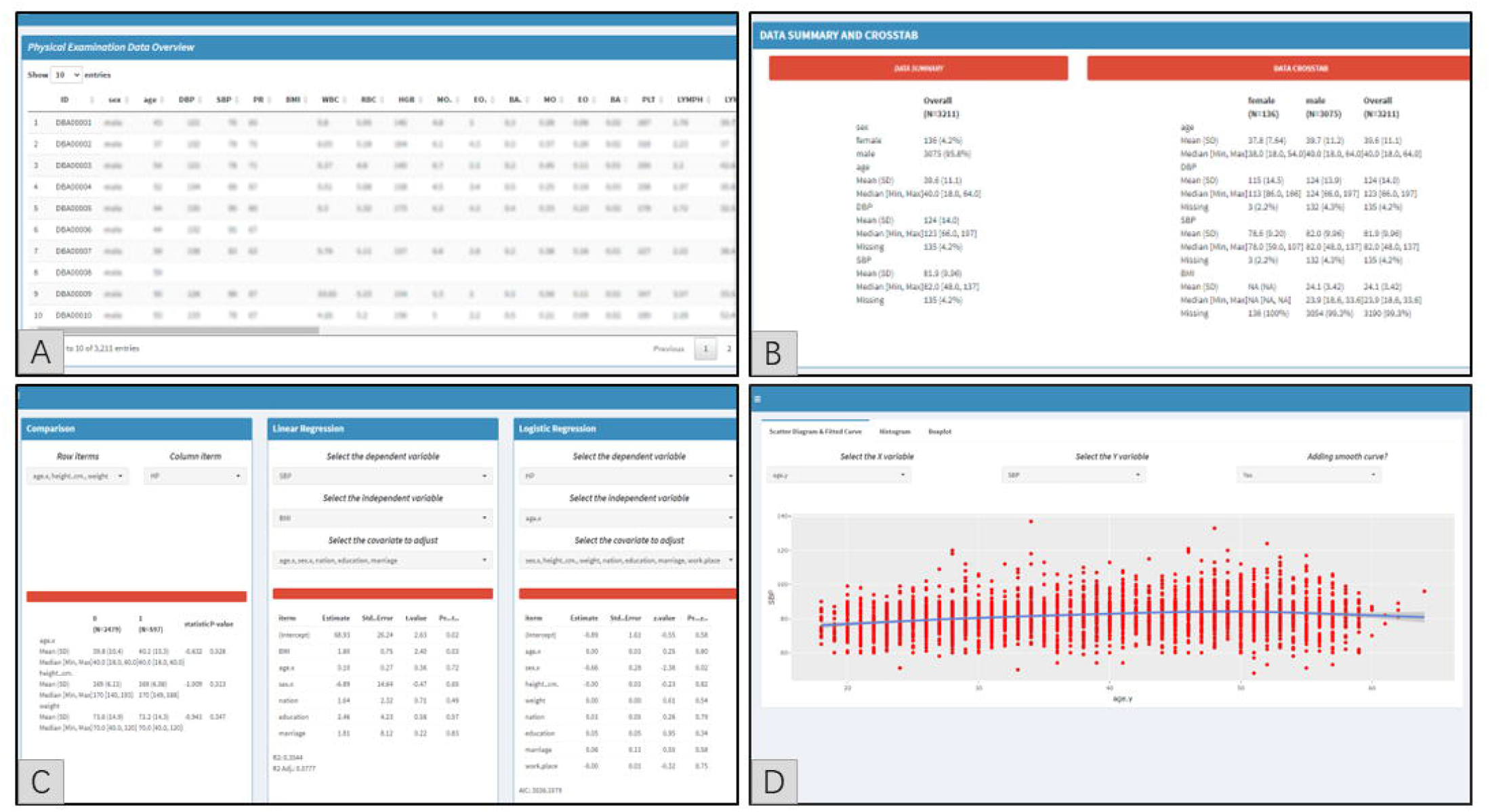

## 4. Discussion

With the development of China’s maritime trade, the number of shipping related workers increase sustainingly, however, the attention focusing on their occupational health problems is not enough. Therefore, in present study an epidemiological investigation for port employees in Shenzhen was implemented and a dataset management system was built, aiming at providing basic data for knowing about the health condition of port employees, and exploring the relationship between occupational hazards exposure and health.

Descriptive analysis was used to demonstrate basic characteristic of survey data in this study, among these participants, the jobs require a lot of physical labor only accounted for 11.9%∼26.4%, which indicated that with equipment upgrading, numerous intelligent equipment and machines have replaced the simple, repetitive and strenuous physical jobs in the port, which reduces the physical dependence to a certain extent. Although the manual labor level has decreased, the level of mental labor has increased, therefore, the occupational stress and health problems according to mechanical vibration and noise, prolonged working posture and concentration, and other occupational hazards may gradually be more obvious^19^. The occupational disease spectrum of port employees may change because of the change of working practices, this change suggests that we should cause our attention

Analysis result indicates that missing data were mainly concentrated in the physical examination dataset, such as motor and neurological function examination items, because these items were fill in the results by professional physicians after examining and diagnosing, if the result is normal (negative), leaving the item blank by default, and if the result is abnormal, the item will be fill in. To be respectful of the authenticity and integrity of the dataset, the original dataset is retained without adding or deleting the null values. Oppositely, in the questionnaire dataset not missing values are observed, mainly due to the utilization of structured electronic questionnaires. Compared with traditional paper questionnaire, anti-missing value mechanism can be set up to avoid missing value: users must fill in the current question before answer the next question. It is useful for improving the completion rate of questionnaire, and evaluating the quality of the dataset. Cronbach’s Alpha coefficient was first proposed by Lee Cronbach^20^, which can assess the internal consistency of the questionnaire by calculating the correlation coefficient between questions. This coefficient has been used widely, especially in survey study^21, 22^. In this study, the total Cronbach’s alpha confidence of ERIQ and NHLMQ was 0.808, which suggests that the dataset is reliable and can be used to further analysis.

The OHSPEIMS provides a simple, fast, free solution for constructing a private, highly customized database management system. Shiny is often used to build interactive Web apps, and more and more apps are being published online to simplify some complex analysis processes and data visualization, such as genomic analysis^23^, proteomic analysis^24^, transcriptome analysis^25^, prediction model^26^, complex model^27^ and visualization system for COVID-19 pandemic^27^. Hence, it is entirely possible to build a customized dataset management system using Shiny. In addition, R software contains a huge number of packages to facilitate the user to complete more functions, and those packages are developed and updated continuously through the concerted efforts of researchers around the world, which provides convenience for upgrading and improving the database system. In this study, OHSPEIMS is a rough framework needing optimization and update according to the job requirements in the future, for example, adding ‘glmnet’ package to conduce more complex linear or nonlinear regression analysis^29^; using the ‘bkmr’ package can assess the combined exposure effects of multiple occupational hazards^30, 31^; and predicting the occupational health outcome by utilizing machine learning algorithm via ‘mlr’ package^32, 33^.

However, potential limitation of this study needing to point out: First, the investigation is cross-sectional and retrospective design, which exists the challenges in information bias, insufficient evidence and causal inference. Second, paucity of environmental monitoring data failure to assess the occupational exposure directly or indirectly. Finally, the function of OHSPEIMS is relatively simple, it is difficult to achieve further analysis. Therefore, long-term follow-up survey, environmental monitoring and system optimization are necessary in further studies.

## 5. Conclusion

OHSPE is a reliable survey and the OHSPEIMS can be used to manage and analyze the dataset, however, further optimization and improvement are still required.

## Data Availability

Not applicable.

## Acknowledgments

Not applicable.

## Author contributions

Conception and design: Jinlin Wang and Chunbao Mo; Acquisition of data: Jinlin Wang, Jinlin Wang, Dafeng Lin, Jie Situ and Ming Zhang; Analysis and interpretation of data: Chunbao Mo; Writing, review, and/or revision of the manuscript: Jinlin Wang and Chunbao Mo; Administrative, technical, or material support: Naixing Zhang; All authors approved the final manuscript.

## Availability of data and materials

Not applicable.

